# Sensorimotor control of object manipulation following middle cerebral artery (MCA) stroke

**DOI:** 10.1101/2022.08.04.22278444

**Authors:** Kelene A. Fercho, Jamie L. Scholl, KC Bikash, Taylor J. Bosch, Lee A. Baugh

## Abstract

Current bedside diagnostics used for the assessment of the loss of hand function post-stroke examine limited aspects of motor performance. Further, they are not sensitive to subtle changes that can cause deficits in everyday object manipulation tasks. Efficiently lifting an object is a complex neurological event which entails a prediction of required forces based on intrinsic features of the object (sensorimotor integration), short-term updates in the forces required to lift objects that are poorly predicted (sensorimotor memory), as well as the ability to modulate distal fingertip forces. Unfortunately, this complexity is not represented by the existing assessment tools used in clinics for both diagnostic and rehabilitative purposes. The presented research examined these three critical components of skilled object manipulation—production of finely graded muscular forces, sensorimotor integration, and sensorimotor memory—in a heterogeneous population of 60 chronic, unilateral middle cerebral artery stroke participants. Performance was compared to age-matched control participants in each of the three tasks. To examine control of distal fingertip forces, a force-matching task was utilized. To examine sensorimotor integration, participants were presented with familiar objects –large wood or brass blocks—following lifting trials of small and medium sized blocks from the same size-weight families. To accurately predict the weight of the larger blocks, sensorimotor integration of object size and apparent material is required during the first lifts of the large blocks. To examine sensorimotor memory, participants were required to lift a series of size-weight blocks of different colors. One color signified an inverse size-weight relationship that required the modification of short-term sensorimotor memory to efficiently lift. Most post-stroke participants performed below control levels in at least one of the tasks. Importantly, post-stroke participants presented with several different combinations of deficits in each of the tasks performed. The presented research demonstrates MCA stroke patients may have deficits in one or more components required for the successful manipulation of hand-held objects. Further, this information may be used in future studies to aid efforts that target rehabilitation regimens to a stroke survivor’s specific pattern of deficits.

## 1. Introduction

With an estimated 7.0 million Americans over the age of 20 having had a stroke and more than 795,000 experiencing a first or recurrent stroke each year, stroke is the leading cause of preventable disability in the United States (Virani, et al., 2020). Upper-extremity impairments including fine motor control often occur following the most common ischemic stroke, that which affects the middle cerebral artery (MCA). These impairments are largely due to the role of the MCA in supplying blood to the lateral surfaces of the brain. Specifically, the action of skilled precision grip and lift represents a diverse network of brain regions including significant subdivisions of the frontal and parietal lobes required for processing different features of skilled hand and arm movements (Errante, et al., 2021). Further, when lifting familiar objects, healthy adults predictively program the required fingertip and lifting forces based on the physical properties of the object such as mass and friction. These properties are largely learned from interactions with similar objects within the environment (Cole & Rotella, 2002; Gordon, et al., 1993; Johansson & Westling, 1988) with the ability to accurately predict the weight of the lifted object being essential to efficient performance (Flanagan, et al., 2006; Wolpert & Flanagan, 2001). For example, when lifting an object off a surface like a tabletop, people will smoothly increase vertical load force to a level that is slightly higher than the weight of the object, allowing the object to be accelerated upward during the lift. When this prediction is well-matched to the properties of the object, such as its weight, the object is efficiently lifted. Therefore, effective feed-forward motor control strategies allow us to perform fast and accurate voluntary grasps that are not possible when only feedback mechanisms are available.

Upper-extremity dysfunctions commonly observed following MCA stroke, even when mild, can have debilitating effects on activities of daily living, including object manipulation tasks (Eidenmüller, et al., 2014; Lindberg, et al., 2012; Nowak & Hermsdörfer, 2003). In turn, these dysfunctions substantially reduce overall quality of life (Ekstrand, et al., 2016) and result in avoidance of interacting with objects that require a fine-tuning of grip forces (i.e., fragile objects). Additionally, patients experience increased fatigue from using exaggerated manipulation forces in common everyday manipulation tasks (Hermsdörfer, et al., 2003).

It has been previously shown that damage to components of the cortical and subcortical network that support predictive force control during object manipulation can result in reliance on a compensatory feedback response that is less efficient, more time consuming, and utilizes increased safety margins, lowering the overall ability to interact with one’s surroundings in a skillful manner (Gordon, et al., 1997; Hermsdörfer, et al., 2003; Nowak, et al., 2003; Quaney, et al., 2005). Further, previous research has demonstrated that following a unilateral MCA stroke, patients often display prominent deficits in this ability, even when utilizing the ipsilesional “preserved” hand, providing strong evidence that regions higher than basic motor and premotor regions are involved (Quaney, et al., 2005). It has been suggested that these areas support three critical components of skilled object manipulations: 1) An ability to produce finely graded muscular forces; 2) The ability to adequately perform sensorimotor integration; and 3) The acquisition of motor memory (Quaney, et al., 2010). First, the ability to produce finely graded, low intensity forces from the distal muscles is often affected by the disruption of neural pathways related to the injury (Jo, et al., 2016; Li, et al., 2003; Roh, et al., 2015). Next, the ability to integrate ascending somatosensory information within cortical regions could be impaired, resulting in an inability to combine information about object shape, texture, and weight into a useful percept for future interactions with that object (Edwards, et al., 2019; Wolpert, et al., 1998). Lastly, even if distal muscles can provide the fine-scale force required for object manipulation and somatosensory information is being effectively combined, storing object representations within sensorimotor memory for future interactions may be deficient.

Although each of these required facets of skilled object manipulation is known, there has been little attention to how each of these abilities is affected following MCA stroke. Based on the territories irrigated by the MCA and the known neural correlates of each of these tasks (for a review see Errante, et al., 2021), a single stroke survivor may have impairments in one or more of these systems. In this study, we sought to determine the pattern of deficits in each of these abilities in a heterogeneous population of 60 chronic unilateral MCA stroke patients with mild upper-extremity deficits using tasks designed to assess the distal production of forces in the dominant hand, sensorimotor integration, and sensorimotor memory. We hypothesized that these abilities are dissociable, with stroke participants with upper-extremity dysfunction having deficits in one or more of these abilities and that these impairments could affect both the affected contralesional and “preserved” ipsilesional hand.

Since current bedside assessment tools do not represent the predicted complexity of possible deficits in skilled object manipulation, the information from this study may increase our ability to identify each of these unique abilities, ultimately aiding in future efforts to target rehabilitation regimens to a stroke survivor’s specific pattern of deficits.

## 2. MATERIALS AND METHODS

### 2.1 Participants

All procedures were approved by the Institutional Review Board of the University of South Dakota, and were compensated $20 USD per hour for their time. Sixty right-handed adults with chronic unilateral MCA stroke (29 Female; Mean Age 65 years old; Average time since stroke 1335 days; 18 left hemisphere damage [LHD], mean Action Reach Arm Test [ARAT] score of dominant right hand = 57), and 48 healthy right-handed adults (30 Female, Mean Age 76 years old) participated in a series of three experiments. Within both the control and post-stroke groups, there were several participants that did not complete all tasks due to time (a session was limited to three hours), fatigue, task difficulty, or disinterest. Specific numbers of participants that completed each of the tasks are provided within the results section.

Participants completed the tasks with their dominant right hand. All stroke survivors had low levels of dysfunction in their affected upper limb with mild paresis represented by a minimum ARAT score greater than 50 (Lyle, 1981). The patients were selected based on the presence of a single unilateral hemisphere MCA stroke confirmed by radiologist interpretation of the computed tomography or magnetic resonance imaging scan, as well as a clinical examination showing deficits of fine motor control within the upper extremities without sensory deficits (as assessed by Semmes-Winstein monofilament testing values of 3.61 – 4.65g). None of the patients exhibited pronounced spasticity of the affected upper extremity and none had any other history or clinical signs of other neurological diseases as assessed by their practicing neurologist. Stroke participants were excluded if they showed symptoms of receptive aphasia, apraxia, or cognitive deficits demonstrated through the Folstein Mini-Mental Status Exam (Folstein, et al., 1975), medical chart review, and/or physician evaluation. The stroke event must have occurred more than 6 months prior to participation in the study. Handedness was assessed for all participants via a modified Edinburgh handedness inventory (Oldfield, 1971).

### 2.2 Apparatus and Procedure

Task order was randomized across participants to ensure that order effects, especially fatigue, did not affect one task more than the others.

#### 2.2.1 Force Matching Task

A visually guided ramp-and-hold force matching task was used in which participants matched a target force by generating isometric precision grip forces (Lindberg, et al., 2009). Participants were seated in a chair and grasped a handle instrumented with a force/torque sensor (Nano 17 F/T sensors; ATI Industrial Automation, Garner, NC) sampling at 1000Hz. The forearm was flexed at roughly 45 degrees, and the elbow was positioned on the table to ensure a steady posture throughout the experiment. Participants were required to perform a visuomotor step-tracking task by creating isometric force between the thumb and index finger. Measured forces and the target force were displayed to the participants in real-time on a 15” LCD monitor placed at the midline (Figures 1A and 1B). Required forces ranged between 0 and 30% of a participant’s maximum voluntary contraction (MVC) rate. Isometric forces were filtered online with an 8Hz, 2^nd^ order Butterworth filter to remove the participants’ natural physiological tremor before being digitized for display purposes. Instructions were given verbally, with participants being asked to hold the instrumented manipulandum in a precision grip, apply the maximum amount of isometric force possible, and maintain that force for five seconds to measure the MVC. Immediately following this procedure, participants completed 150 trials of the visuomotor tracking task by exerting isometric force between the thumb and index finger. The computer monitor displayed two vertical scales representing both the instantaneous force exerted on the sensor and the target force at all times. Target forces were calculated and randomly presented at 5, 10, 15, 20, 25, and 30% of MVC. Participants were instructed to match the target force as closely as possible until trial completion defined as a generated force within 0.5N of the target force for a minimum of 1s, upon which the next target force was immediately displayed. The entire task took no longer than 20 minutes to complete. To avoid changes in hand and grip positioning, participants were monitored and instructed to maintain their index finger and thumb on the force transducers. All participants were familiarized with the task before testing.

**Figure 1.**
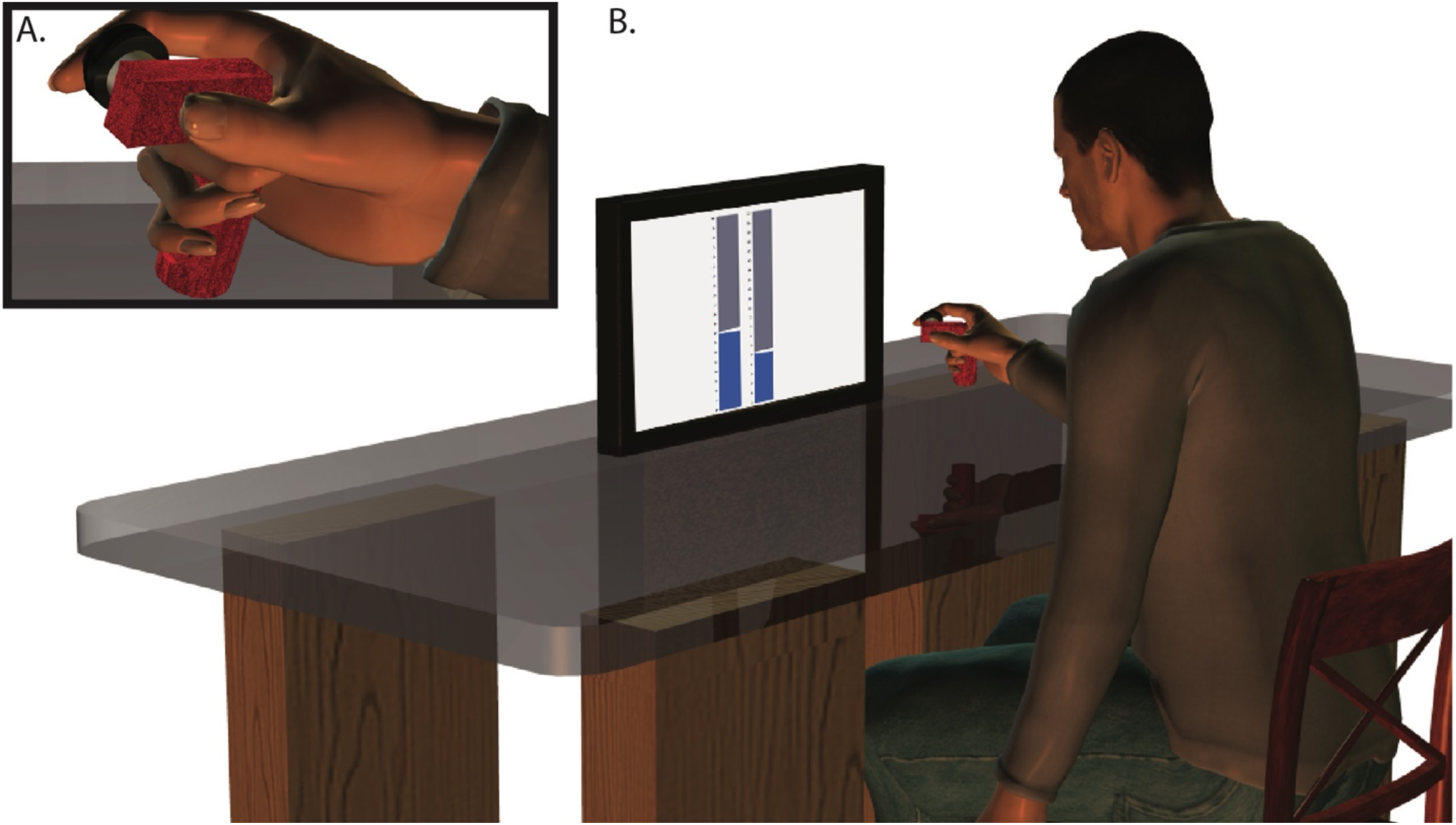
Force Matching Task. Participants applied force to the manipulandum using a precision grip (A). Participants were required to match the pincher force requested using the hand-held manipulandum. Both visual and numerical representations of the required and current force levels were presented to the participant via an LCD screen (B).

#### 2.2.2 General Object Lifting Tasks

The apparatus used for the object lifting tasks is pictured in Figure 2A. For each trial, the participant lifted an object from a platform equipped with a force sensor (Nano 17 F/T sensor; ATI Industrial Automation, Garner, NC) sampling at 500Hz and then placed it back on the tabletop in the same location. Participants were prompted via a 15” LCD screen to lift the object using their dominant hand. In all lifting trials, participants wore LCD shutter glasses (Plato Technologies, Toronto, Ontario, Canada) that blocked visual cues to object weight that may have been provided by observing the experimenter place each block during the inter-trial intervals. Participants received verbal instructions and a demonstration by the experimenter showing them how to perform the lifting motion. In all cases, an auditory tone (500Hz, 1sec) indicated when the participant was to begin the lift and coincided with the shutter glasses turning translucent. A second, lower-pitched tone (250Hz, 1sec) signaled when the participant should place the object back down. At the end of each lifting trial, the shutter glasses turned opaque.

**Figure 2.**
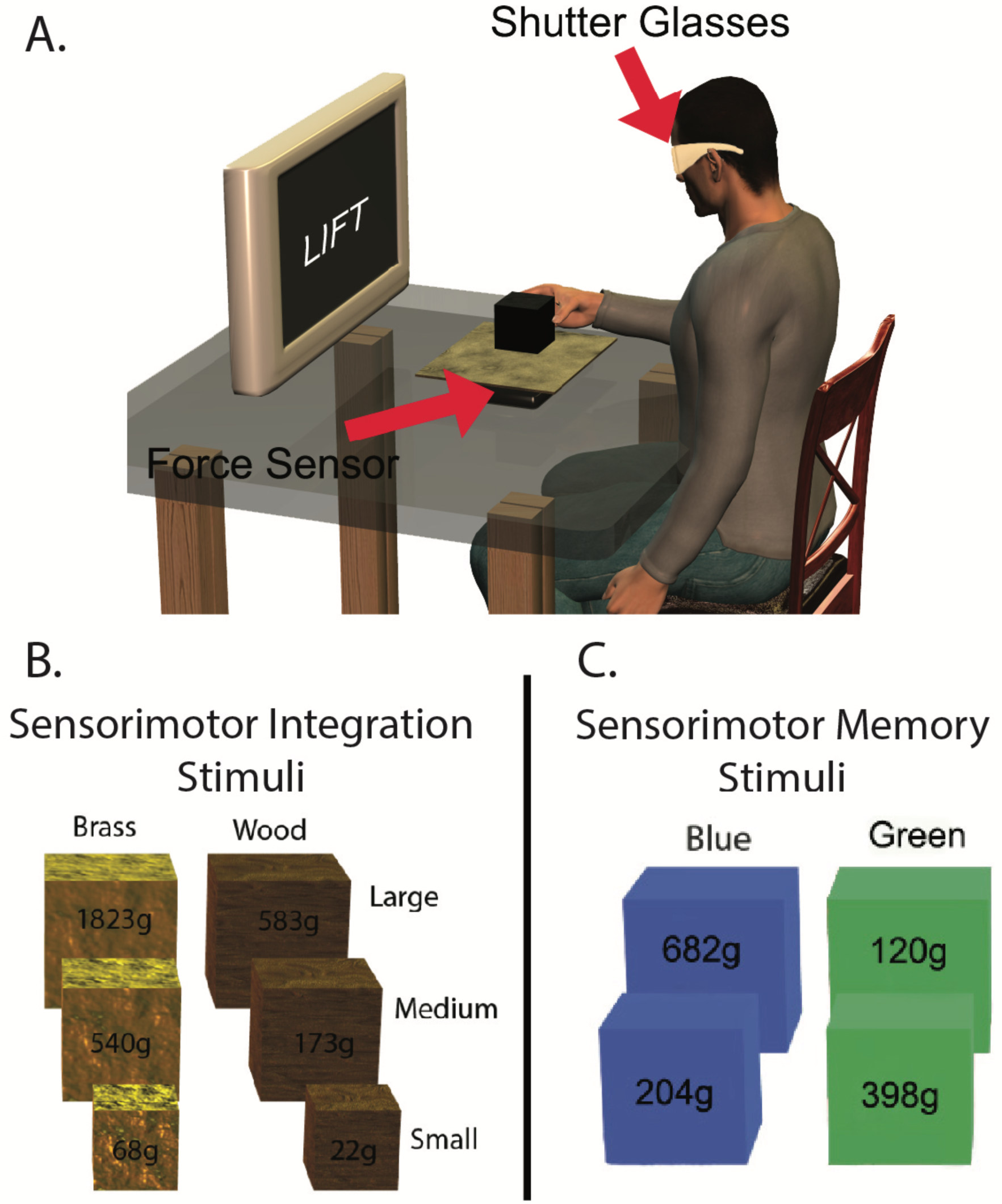
Sensorimotor Integration and Sensorimotor Memory Tasks. Participants lifted the presented block off a platform equipped with Force/Torque sensors. An LCD screen was used to prompt the lifting behavior (A). In the sensorimotor integration task, participants lifted each of the small and medium brass and wood blocks 60 times (120 total lifts), and the large brass and wood blocks 20 times each for a total of 140 lifts (B). Within the sensorimotor memory task, participants lifted each of the inversely weighted blue and green blocks 20 times for a total of 80 lifts (C).

#### 2.2.4 Sensorimotor Integration Task

To assess sensorimotor integration abilities, large wood or brass blocks, representing familiar objects, were presented to participants following trials of small and medium sized blocks from the same material-weight families. The blocks consisted of 3 matched material blocks increasing both in size and weight, with brass: small (3cm^3^, 84g), medium (4.5cm^3^, 244g), and large (6cm^3^, 568g), and wood: small (3cm^3^, 26g), medium (4.5cm^3^, 96g), and large (6cm^3^, 182g) (See Figure 2B). To accurately predict the initial weight of the large blocks, initial integration of object size and apparent material is required. Participants lifted the small and medium blocks 60 times each (120 total lifts) and the large blocks 10 times each (20 lifts) for a total of 140 lifts. Participants received instruction and viewed a demonstration on how to perform the lifting motion. The task consisted of lifting the object approximately 2cm off the tabletop, pausing while holding the object for 1s, and then returning the object to the tabletop in its original position.

#### 2.2.4 Sensorimotor Memory Task

To assess sensorimotor memory, a similar procedure to the sensorimotor integration task previously described was employed. Specifically, participants were required to lift a series of blocks including those with an inversed size-weight relationship. Size-weight matched blocks were blue, with medium (6cm^3^, 204g) and large (9cm^3^, 682g) sizes presented to participants (See Figure 2C). Inverse blocks of green were created with an inverse size-weight relationship, with medium blocks (6cm^3^, 398g) weighing significantly more than large blocks (9cm^3^, 120g). Therefore, to accurately predict the weight of such blocks, sensorimotor memory must be used to recall the relationship between object color and the size-weight mapping of the object family (Flanagan, et al., 2001). Participants lifted each block 20 times (total 80 lifts). To ensure the last lifts were not simply a repeat of successful lifting forces from the previous trial, order was pseudo-randomized with the constraint that the last lift of each block could not follow a lift of the same block in the previous trial.

### 2.3 Data Analysis

#### 2.3.1 Force Matching Task

Raw force signals were low pass filtered using a 4^th^ order, zero-phase lag Butterworth filter with a cut-off frequency of 14Hz offline. The final hold phase start time was calculated for each trial as the time taken to successfully generate an isometric force within 0.5N of the target force and hold the generated force within that range for 1s (See Figure 3). Therefore, this timed metric reflected both errors in precision and speed in distal force generation. Following omnibus repeated-measures ANOVA (rm-ANOVA) with force level (Low vs. High) as the repeated variable, and Group (Control, right hemisphere damage [RHD], left hemisphere damage [LHD]) as a between subjects factor to examine the overall effect of stroke on task performance, hold phase start time for the control participants was used across all trials to create a mean and standard deviation. This mean and standard deviation allowed for the conversion of individual stroke participant performance to Z-Scores relative to control performance. This approach allowed for a direct comparison of each individual stroke participant to control data.

**Figure 3.**
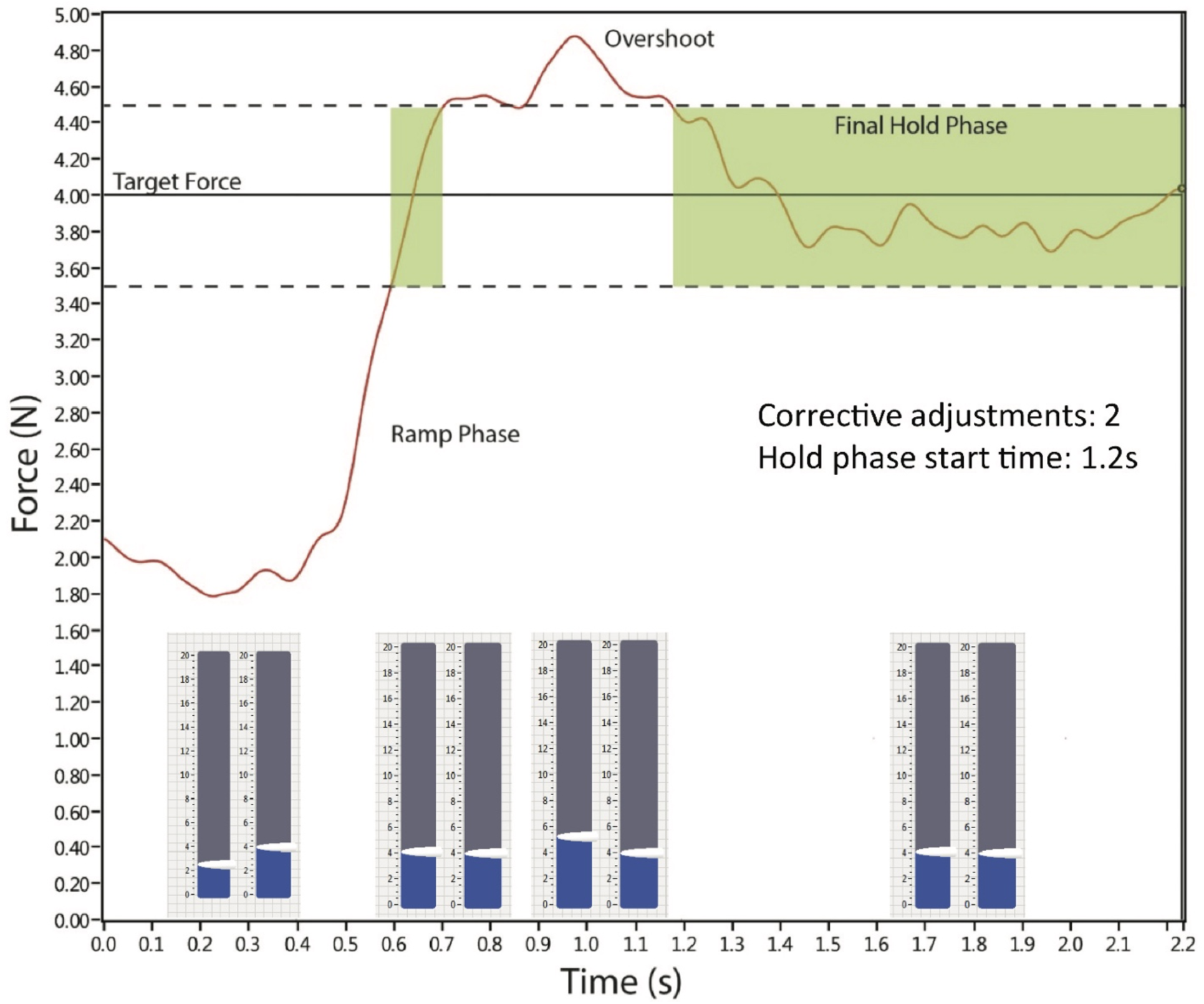
Force Matching Task Analysis. Raw force signals were low pass filtered using a 4^th^ order, zero-phase lag Butterworth filter with a cut-off frequency of 14Hz offline. The final hold phase start time was calculated for each trial as the time taken to successfully generate an isometric force within 0.5N of the target force and hold the generated force within that range for 1s.

#### 2.3.2 Object Lifting Tasks

Raw force signals were low-pass filtered using a 4^th^ order, zero-phase lag Butterworth filter with a cutoff frequency of 14Hz. A signal representing the vertical force applied to the object by the hand (i.e., the vertical lifting or load force) was obtained by subtracting the vertical force that accounted for the object weight when fully supported by the transducer from the recorded signal. The processed signal was then differentiated with respect to time using a 1st order central difference equation to obtain the rate of change in load force, or load-force rate. For each lift, we determined the load-phase duration (See Figure 4). The start of the load phase was defined as the time when load force first exceeded 0.2N. Thus, the first peak in load-force rate (a maxima followed by a decrease) had to occur after the load force exceeded 0.2N. A threshold of 0.2N was selected because load-force values early in the lift below this value would not be attributable to obvious attempts to lift the object but rather to initial finger placement on the block. The end of the load phase was defined as the time, just before object lift-off, when load force reached within 0.2N of the object weight. Use of load phase duration as a measurement of lifting efficiency is possible as people normalize load phase durations across object weight by scaling the load force rate to the expected weight of the object. Therefore, load phase duration is reflective of the participants’ predicted weight of the object. Following omnibus rm-ANOVAs to examine the effects of either left or right hemisphere stroke on object lifting performance, control participant data was used to create a mean and standard deviation to allow for the conversion of stroke participant performance to Z-scores, allowing for a direct comparison of each individual stroke participant to the healthy control participants’ performance.

**Figure 4.**
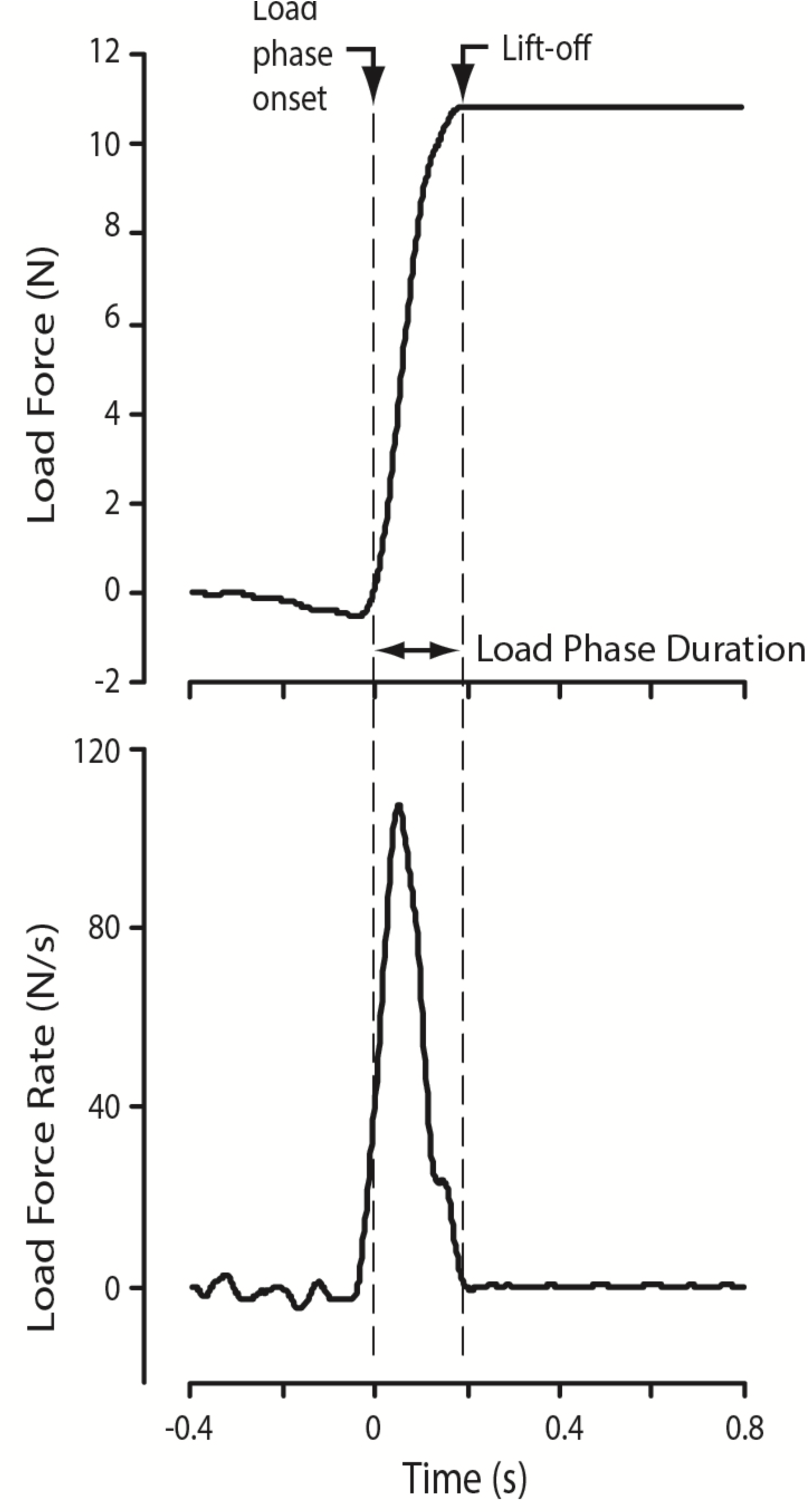
Object Lifting Task. Load phase duration was calculated offline post-experiment on a trial-by-trial basis. The start of the load phase was defined as the time when load force first exceeded 0.2N. The load phase ended at object lift-off. Therefore, load phase duration was the defined as the time between load phase onset and object lift-off.

#### 2.3.3 Sensorimotor Integration Task

Data analysis focused on the load phase durations of the first three and last three lifts of the large brass blocks using a rmANOVA with lift (first three vs. last three) as the repeated factor and Group (RHD vs. LHD vs. Control) as a between-subjects variable. The first three lifts were examined to prevent the influence of sensorimotor memory on subsequent lifts, whereas the last three lifts were examined to see whether participants were able to perform within normal ranges following direct experience with the large brass blocks. Based on the experience participants had lifting the small and medium sized blocks, sensorimotor integration should allow participants to use information about the novel large blocks (size and apparent material) to adequately program lifting forces when first lifting the large brass blocks (Baugh, et al., 2012).

#### 2.3.4 Sensorimotor Memory Task

Data analysis focused on the last three lifts of each block to allow the most exposure to each of the objects. Further, to prevent the influence of previous trials, these were the only trials where the object lifted did not follow an identical object used in the previous lift.

## 3. RESULTS

Final participant numbers for each of the tasks can be seen in Table 1. In brief, 23 participants completed all three tasks with their dominant hand. An additional 24 participants completed two of the three tasks, with 13 stroke participants completing only one of the tasks.

**Table 1.**
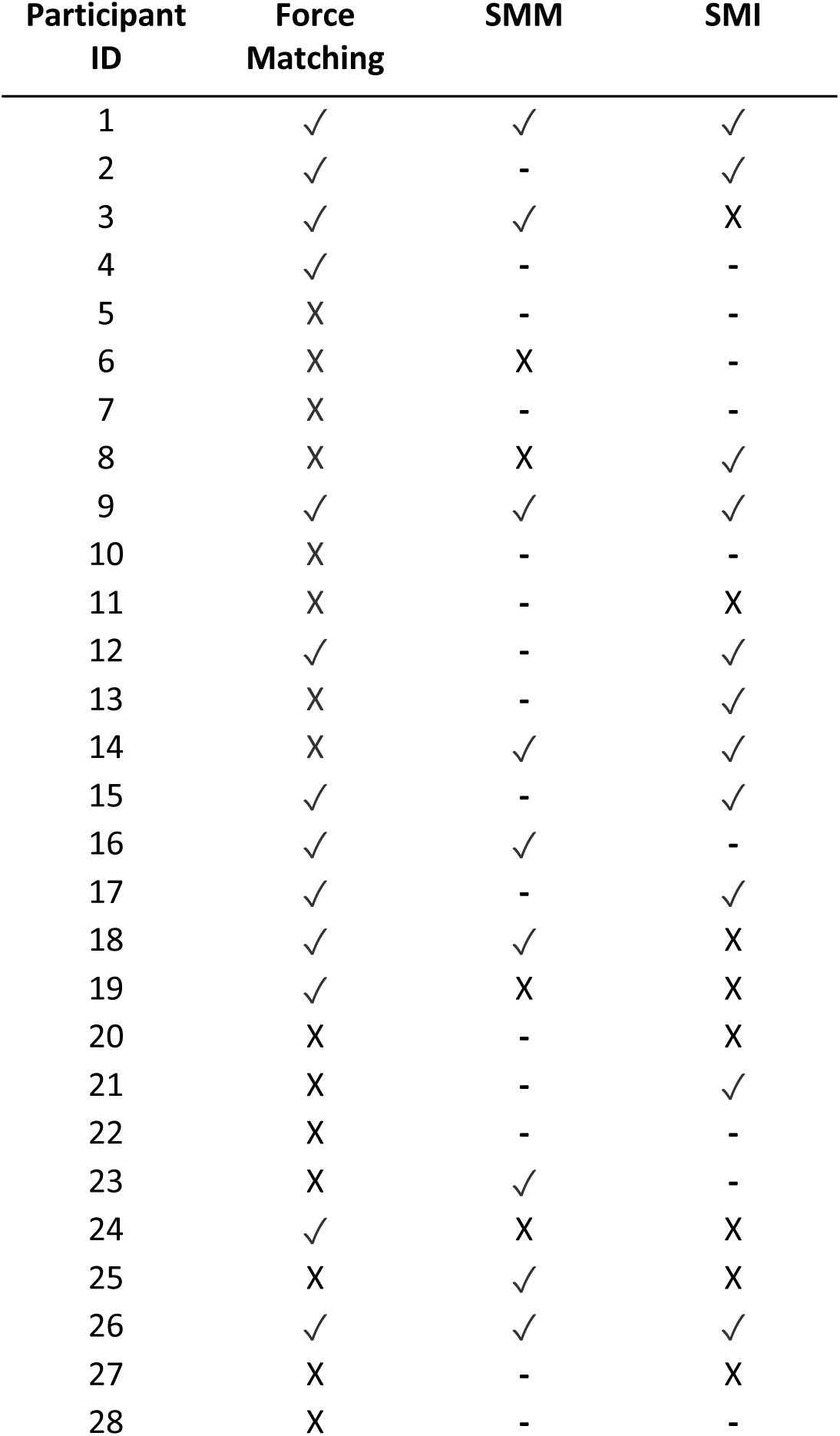
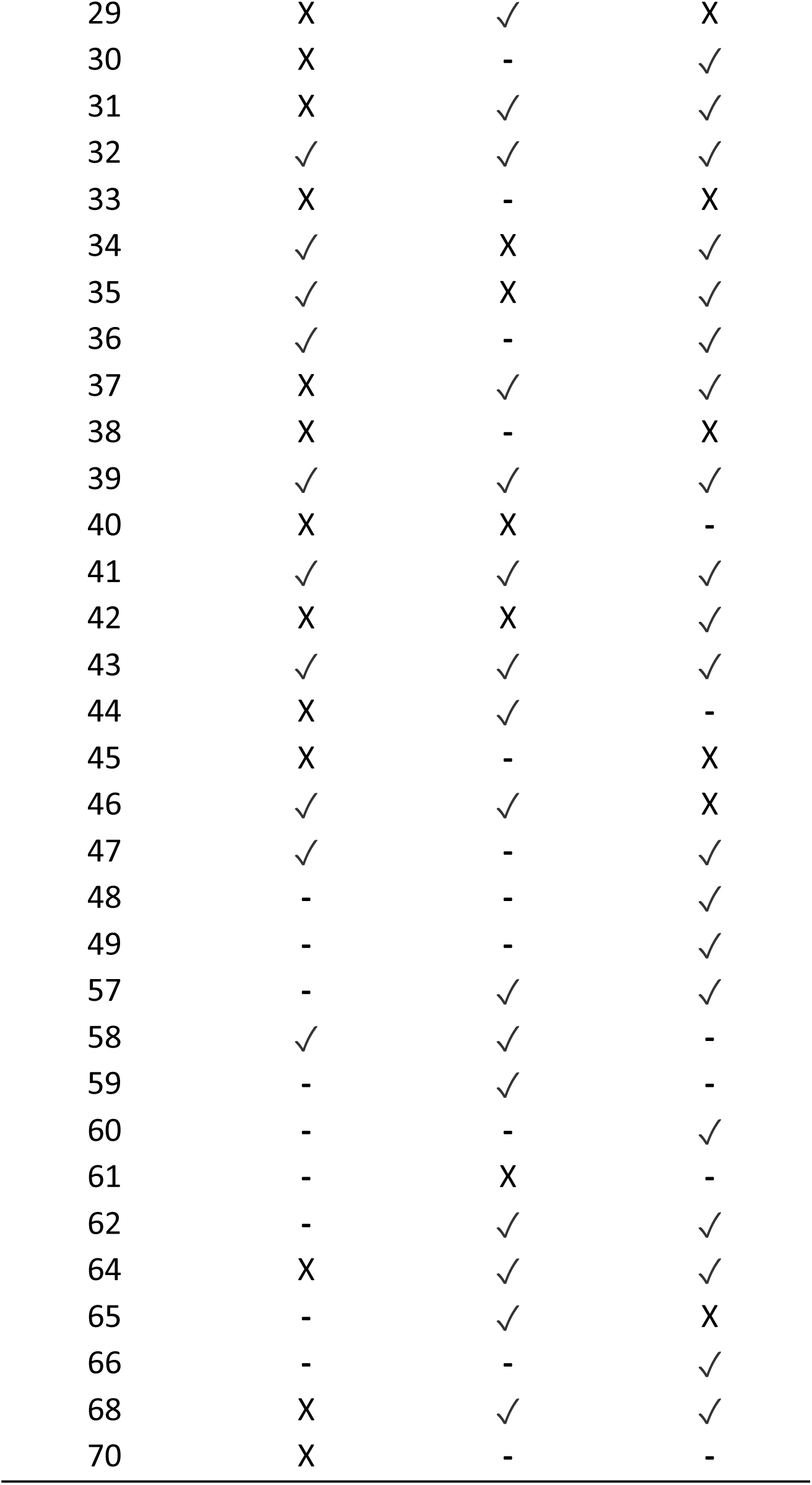
Final Stroke Participant Numbers. Twenty-three participants completed all three tasks with their dominant hand. An additional 24 participants completed two of the three tasks, with 13 stroke participants completing only one of the tasks for a total of 60 participant’s data. A checkmark denotes performance within the normal range, an ‘X’ denotes performance outside of the normal range and the ‘-’ denotes the participant did not complete the task. SMM = Sensorimotor Memory, SMI = Sensorimotor Integration.

| Participant ID | Force Matching | SMM | SMI |
| --- | --- | --- | --- |
| 1 | ✓ | ✓ | ✓ |
| 2 | ✓ | - | ✓ |
| 3 | ✓ | ✓ | X |
| 4 | ✓ | - | - |
| 5 | X | - | - |
| 6 | X | X | - |
| 7 | X | - | - |
| 8 | X | X | ✓ |
| 9 | ✓ | ✓ | ✓ |
| 10 | X | - | - |
| 11 | X | - | X |
| 12 | ✓ | - | ✓ |
| 13 | X | - | ✓ |
| 14 | X | ✓ | ✓ |
| 15 | ✓ | - | ✓ |
| 16 | ✓ | ✓ | - |
| 17 | ✓ | - | ✓ |
| 18 | ✓ | ✓ | X |
| 19 | ✓ | X | X |
| 20 | X | - | X |
| 21 | X | - | ✓ |
| 22 | X | - | - |
| 23 | X | ✓ | - |
| 24 | ✓ | X | X |
| 25 | X | ✓ | X |
| 26 | ✓ | ✓ | ✓ |
| 27 | X | - | X |
| 28 | X | - | - |
| 29 | X | ✓ | X |
| 30 | X | - | ✓ |
| 31 | X | ✓ | ✓ |
| 32 | ✓ | ✓ | ✓ |
| 33 | X | - | X |
| 34 | ✓ | X | ✓ |
| 35 | ✓ | X | ✓ |
| 36 | ✓ | - | ✓ |
| 37 | X | ✓ | ✓ |
| 38 | X | - | X |
| 39 | ✓ | ✓ | ✓ |
| 40 | X | X | - |
| 41 | ✓ | ✓ | ✓ |
| 42 | X | X | ✓ |
| 43 | ✓ | ✓ | ✓ |
| 44 | X | ✓ | - |
| 45 | X | - | X |
| 46 | ✓ | ✓ | X |
| 47 | ✓ | - | ✓ |
| 48 | - | - | ✓ |
| 49 | - | - | ✓ |
| 57 | - | ✓ | ✓ |
| 58 | ✓ | ✓ | - |
| 59 | - | ✓ | - |
| 60 | - | - | ✓ |
| 61 | - | X | - |
| 62 | - | ✓ | ✓ |
| 64 | X | ✓ | ✓ |
| 65 | - | ✓ | X |
| 66 | - | - | ✓ |
| 68 | X | ✓ | ✓ |
| 70 | X | - | - |

### 3.1 Force Matching Task

ANOVA revealed a significant main effect of Group (F_(2,71)_ = 3.44, p = .038, ω^2^ = .062). Post-hoc testing revealed a significant difference between the RHD and control group (Mean Difference [MD] = 1688, p = .012, d = .73). The difference between the LHD and control was not statistically significant (MD = 1196, p = .08, d = .517) (See Figure 5A). When examining individual patient performance, 28 participants (12 LHD, 16 RHD) performed outside of the normal control range based on computed z-scores (See Figures 5B and C).

**Figure 5.**
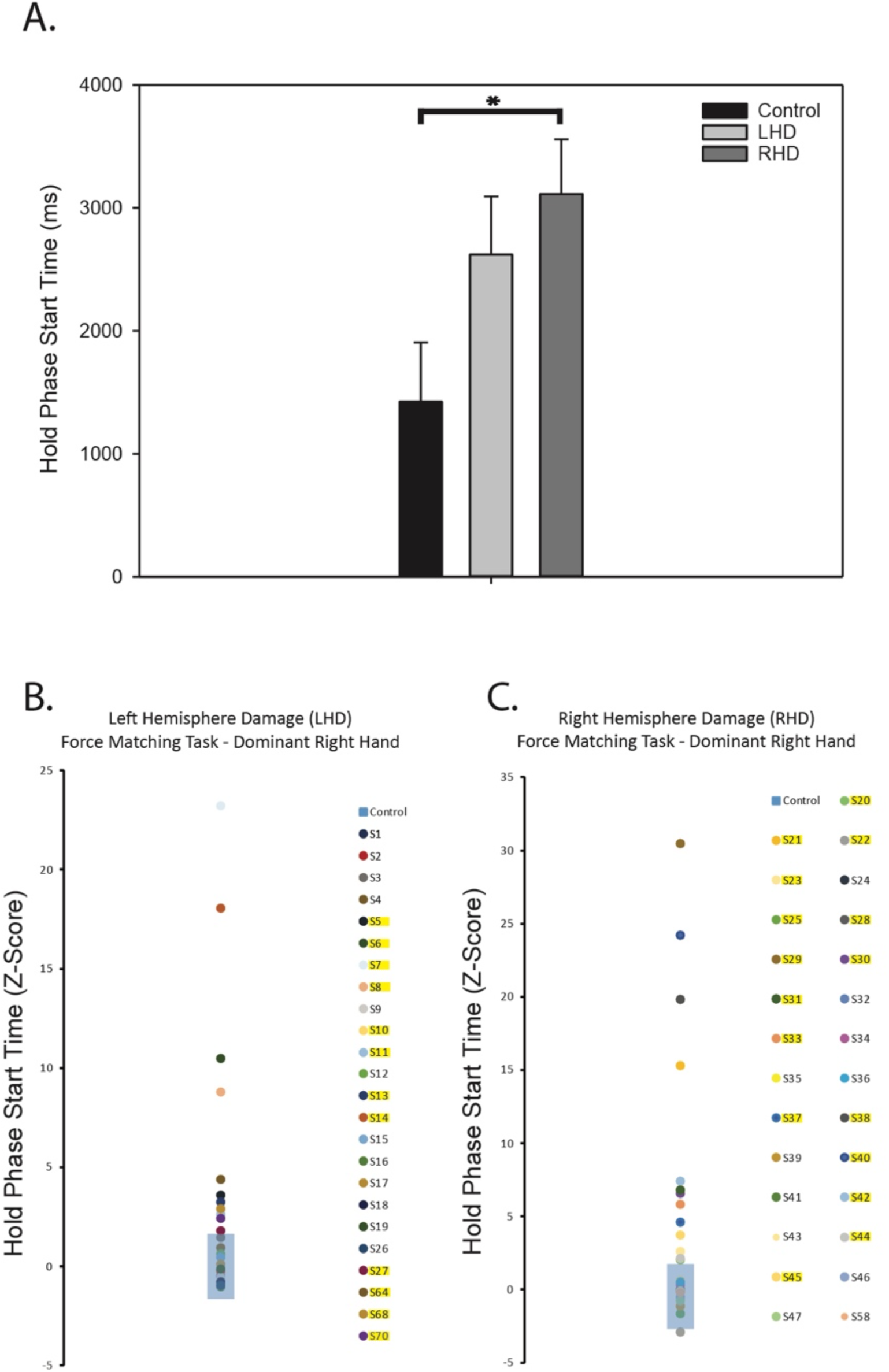
Force Matching Task Results. Overall, stroke participants had greater load-phase start times than controls. Post-hoc testing revealed participants with right-hemisphere damage (RHD) had increased hold phase start times when compared to control participants (A). When examining individual patient Z-scores, compared to control participants, 12 RHD participants performed outside of the control range (B) and 16 left-hemisphere damage (LHD) participants performed outside of the control range, as indicated by the highlighted participant numbers. The blue rectangles represent the healthy control range. *Denotes post-hoc testing p < .05.

### 3.2 Sensorimotor Integration Task

Repeated-measures ANOVA demonstrated a main effect of Time, with early lifts associated with longer load phase durations than later lifts (F_(1,70)_ = 14.48, p < .001, η^2^_p_ = .171). A significant between-subjects effect of Group (F_(2,70)_ = 3.99, p = .023, η^2^_p_ = .102) was also observed, with the stroke participants having longer load phase durations than the control participants. Post-hoc comparisons across group at each time point (Early vs. Late) demonstrated significant differences between the RHD and control group in the early lifts (RHD vs Control – MD = 136 ms, p = .023). No significant difference was observed for the LHD group (LHD vs Control – MD = 130 ms, p = .056) (see Figure 6A). For the last three lifts, post-hoc comparisons revealed a significant difference between the control and LHD group only (MD = 100 ms, p = .005). When examining individual patient performance, 14 participants (5 LHD) fell outside of the normal control range (See Figure 6B and C).

**Figure 6.**
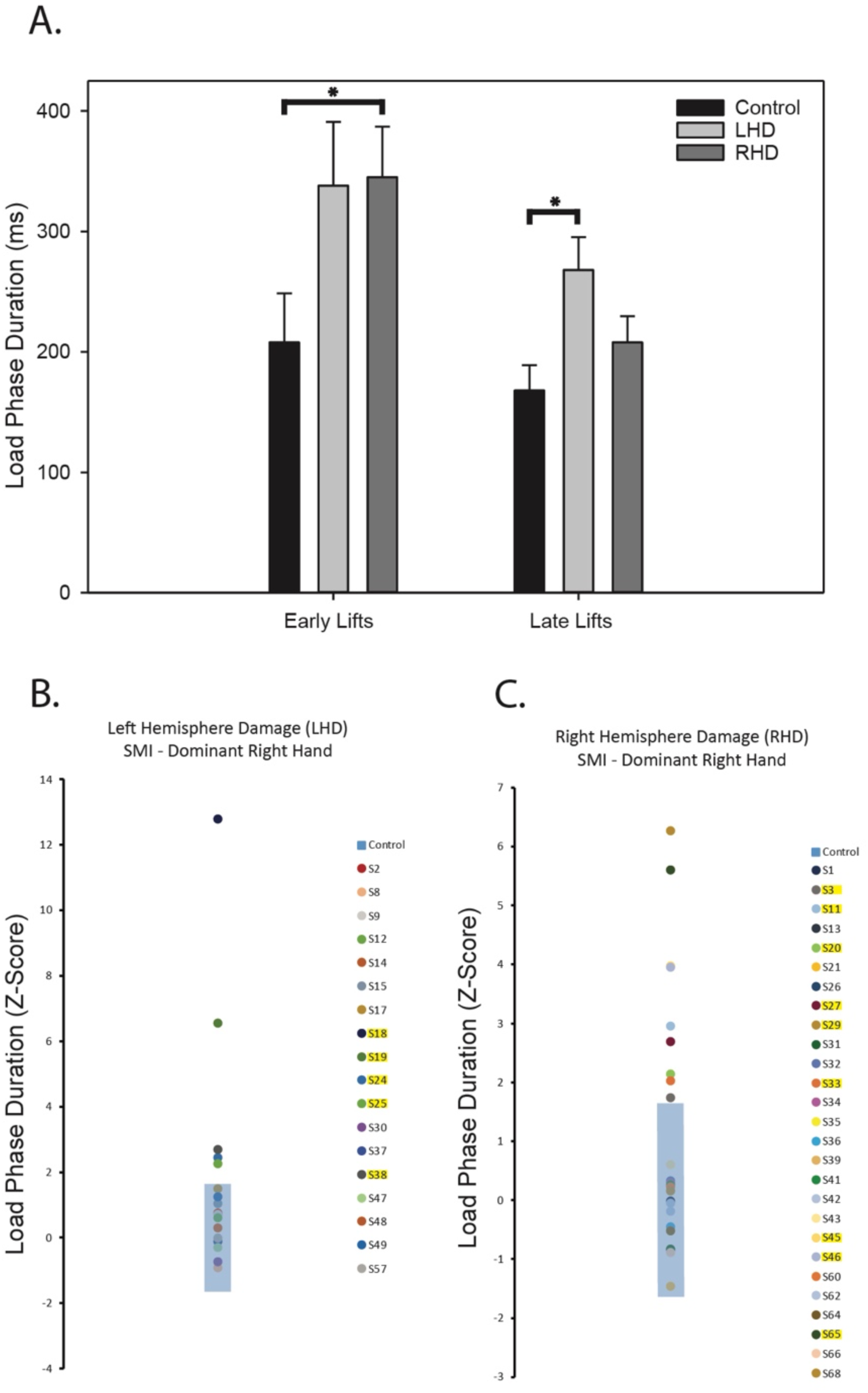
Sensorimotor Integration Results. A main effect of Time was observed, with early lifts being associated with longer load phase durations than later lifts. A significant effect of Group was also observed, with stroke participants having longer load phase durations than control participants. Post-hoc testing confirmed a significant difference between the right-hemisphere damage group and controls in early lifts, and a significant difference between left-hemisphere damaged patients and the controls in later lifts (A). When examining individual patient performance, 5 LHD (B) and 9 RHD (C) patients performed outside of the range of healthy controls (blue rectangles). *Denotes post-hoc testing p < .05.

### 3.3 Sensorimotor Memory Task

Repeated-measures ANOVA demonstrated a main effect of Time (Early vs. Late), with early lifts associated with longer load phase durations than later lifts (F_(1,51)_ = 6.421, p = .014, η^2^_p_ = .112). There was no significant interaction (p > .10) or between-subjects effects (F_(2,51)_ = 3.04, p = .057, η^2^_p_ = .106). Planned comparisons between the control group and the LHD and RHD patient groups found no significant differences in early trials between the controls and the LHD or RHD patients, though it approached significance for the RHD patients (MD = 100 ms, p = .194; MD = 134 ms, p = .050, respectively). However, at later trials there was a significant difference between RHD patients and controls (MD = 73 ms, p = .037) (See Figure 7). When examining individual patient performance, 9 participants (4 LHD) fell outside of the normal control range (See Figure 7A and B).

**Figure 7.**
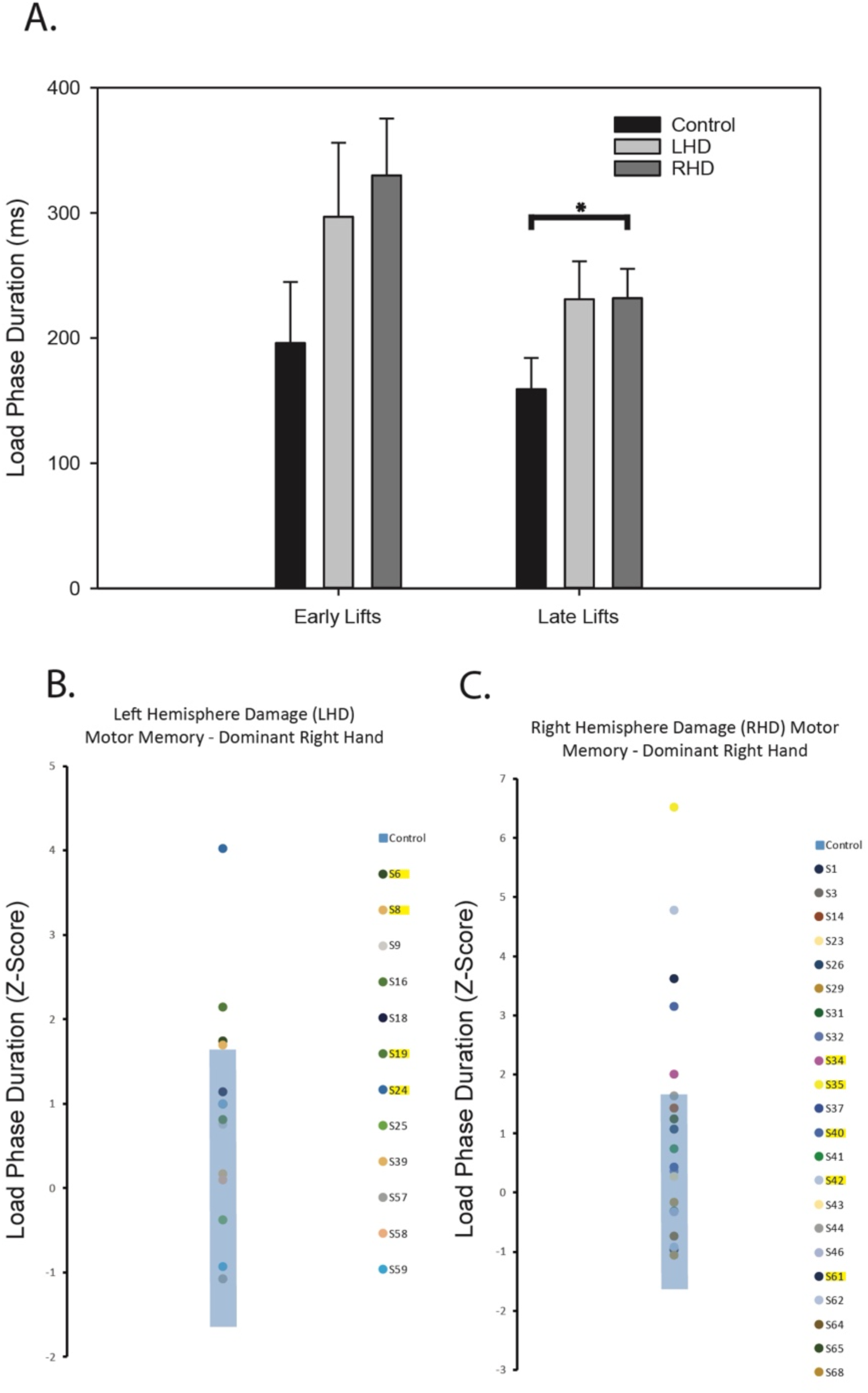
Sensorimotor Memory Results. A main effect of Time demonstrated early lifts had a longer load phase duration than later lifts. Planned comparisons revealed a significant difference between RHD patients and controls (A). Individual patient comparisons revealed 5 RHD (B) and 4 LHD (C) when compared to control (blue rectangles). *Denotes post-hoc testing p < .05.

## 4. DISCUSSION

Interacting with the environment requires a complex series of predictive feed-forward mechanisms, especially when interacting with objects within our surroundings (Buckingham, et al., 2009; Flanagan & Beltzner, 2000; Gordon, et al., 1991; Gordon, et al., 1993; Mon-Williams & Murray, 2000). It is therefore critical to understand how object manipulation abilities are affected by stroke to best develop and assess rehabilitation regimens for stroke survivors. In this study, we sought to determine the pattern of deficits in force matching ability, sensorimotor integration, and sensorimotor memory in a heterogeneous population of right-handed chronic unilateral MCA stroke patients. Each of these abilities is critical to the skillful interaction with objects as part of our day-to-day life (Quaney, et al., 2010). To accomplish this examination, tasks were designed to assess the distal production of forces in the dominant hand, as well as sensorimotor integration abilities and sensorimotor memory during object lifting tasks. Based on diverse neurological systems responsible for each of these abilities (Errante, et al., 2021), we hypothesized these skills would be dissociable following brain damage, with stroke participants with upper-extremity dysfunction having deficits in one or more of these abilities. Further, we predicted that impairment in each of these abilities could be observed in both the affected contralesional and “preserved” ipsilesional hand.

Of the 23 stroke participants that were able to complete all three tasks, 16 scored outside of the range of healthy controls in one or more of the tasks (See Table 1), with 7 scoring within normal control ranges in all three tasks. Of those sixteen, most scored outside of the control range in a single task (N = 10), with the remaining participants (N = 6) scoring outside of the control range in two of the tasks. No participants showed deficits in all 3 tasks. Such a pattern provides strong evidence that stroke survivors may have problems with one or more of these presumably independent systems. Further, patients with both right and left hemisphere damage had difficulty with each of these tasks, a finding that will be discussed in greater detail for each specific task.

### 4.1 Control of Distal Fingertip Forces

Not surprisingly, there was a significant overall effect of Group when looking at the final hold-phase start time. However, this difference was driven more by those in the RHD group than the LHD group. This is a somewhat surprising effect considering the well-known contralateral control of the distal extremities, leading one to expect the contralesional limb to be more affected. All our participants had mild upper-extremity deficits, and therefore the integrity of much of the cortical spinal tract was likely maintained. Therefore, performance deficits in this task are likely not a result of large damage to primary motor cortex or corticospinal tract itself but are more likely a result of damage to supplementary motor areas, as well as the connections between primary and supplementary regions. The impact of unilateral damage to these secondary motor systems is less well understood in terms of laterality of effects.

Of the 51 stroke patients that completed this task, more than half (28) had hold-phase start times outside of the range of healthy age-matched controls. Three patients scored better than age-matched healthy controls, leaving 25 scoring at a lower level of performance. Of these 25 patients, 13 were within the RHD group and 12 were within the LHD group, providing supporting evidence that force production in both the contralesional and ipsilesional limb is affected following unilateral MCA stroke, as has been previously reported (Anderson, et al., 2018; Naik, et al., 2011; Nowak, et al., 2007). When examining performance deficits within the LHD group, difficulty performing the force matching task may arise from several sources resulting in paresis, including damage to both motor and premotor regions (in addition to the disruption of connections between these)(Cash, et al., 2015) and to the white matter pathways ultimately responsible for the innervation of lower motor neurons that allow for direct control of the fingertips (Dum & Strick, 1996; Lang & Schieber, 2003). However, such explanations do not explain deficits in right-handed patients with left-hemisphere damage. Rather, impairments coordinating motor movements, such as what has been reported in previous studies (Hsu, et al., 2018; Schaefer, et al., 2007; Sunderland, et al., 1999), may play a larger role in the deficits observed within the RHD participants. Further work examining the specific nature of these performance deficits is required to elucidate the specific reasons for difficulty with this task in both LHD and RHD participants. As the final hold phase start time is a combined metric, the decreases in performance observed in both groups may be a result of problems with different components of the task.

### 4.2 Sensorimotor Integration

Overall, stroke participants had longer load-phase durations than healthy controls. When comparing the performance of the initial three lifts of the brass block, there was a significant difference between the RHD and control participants, and this result approached significance for the LHD participants. When first lifting the large brass blocks, an increased load phase duration could indicate a problem with sensorimotor integration (taking information about the size and apparent material to predict the weight of the newly encountered block) or with general difficulties in lifting the stimuli. To further explore this possibility, we examined lifting performance during the last three lifts of the large brass blocks. At this point, participants have had ample experience lifting those specific blocks and should rely on additional cues to program their lifting forces in addition to just sensorimotor integration. In the later lifts, there was a significant difference between the LHD and control group, with the RHD group performing at equivalent levels. This suggests that for at least some of the participants in the LHD group, performance may have been impaired for reasons other than difficulties with sensorimotor integration. However, for the RHD group, since their performance improved to control levels, part of their difficulty with the initial lifting is likely a result of deficits in sensorimotor integration, not general lifting ability.

When looking at individual participant performance, this account garners additional support as only 5 of the LHD patients showed performance outside of the healthy control range, whereas 9 with RHD performed at levels below the control group. Even when controlling for the relative number of patients in each group, there were more RHD patients than LHD outside of the control range (33% vs. 27%). These results suggest that the right-hemisphere may have a more prominent role in the sensorimotor integration required for skillfully lifting objects than the left-hemisphere in our group of right-handed patients. This result gains partial support from animal model work, which has demonstrated a right-hemisphere dominance for sensorimotor integration to touch (Grabowski, et al., 1991), as well as electroencephalographic studies showing increased beta power synchronization in right premotor regions during sensorimotor integration processes in a ball catching task (Velasques, et al., 2007).

### 4.3 Sensorimotor Memory

As expected, there was a significant effect of time on load phase durations when examining lifts of the inversely weighted blocks, with most participants improving as they gained experience with this novel relationship between apparent material, object size, and object mass. This effect verifies previous work demonstrating that participants can learn the inverse relationship between size and weight over time. When looking at each of the time points, there were no significant differences between the patient groups (LHD and RHD) and the controls in the early lifts of the blocks (although the RHD group approached a significant difference with a p = .050), suggesting that both patient and controls begin the experiment with inefficient lifting kinematics. Although all load phase durations decreased during the later phases, there was a significant difference between the RHD patient group and the control participants, and this difference approached significant levels for the LHD patients (p = .074).

Once again, when examining individual participant performance, four patients with LHD and 5 with RHD scored outside of the healthy control range. When adjusting for relative numbers of patients within each group, 30% of patients with LHD and 23% of patients with RHD scored outside of the control ranges. This relative increase in the proportion of those with LHD damage scoring outside of the control ranges suggests that although sensorimotor memories are available to both upper extremities (Chang, et al., 2008; Quaney, et al., 2003), they may either be stored within the hemisphere ipsilateral to the arm used to acquire them or within the left hemisphere regardless of hand used. This finding is in concert with recent findings from neuroimaging and neuromodulation studies. In a repetitive transcranial magnetic stimulation study (rTMS), stimulation to dorsal premotor cortex disrupted arbitrary associative memory for weight, whereas rTMS stimulation applied to primary motor cortex disrupted sensorimotor memory during an object lifting task (Chouinard, et al., 2005). In comparison, right inferior parietal cortex, right inferior frontal cortex, and cerebellum all play a role in updating sensorimotor memory representations (Jenmalm, et al., 2006; Schmitz, et al., 2005). Therefore, damage to any one of these unilateral and bilateral systems may result in deficits in updating or utilizing sensorimotor memory resulting in the deficits presently observed.

### 4.4 Limitations

The present study examined patterns of deficits in a heterogeneous population of stroke survivors. Due to the individual variability across participants, and the single-session testing design, many participants were unable to complete all three tasks within the three-hour testing session. Although this may be reflective of specific task difficulty (for example, fewer participants completed the sensorimotor memory task than the sensorimotor integration task), equating a failure to complete the task with task difficulty is problematic. There are many reasons why a participant did not finish a task, and although task order was randomized, general fatigue and other subjective factors may be involved and are uncontrolled, making interpretation difficult. Future studies may offer the most insight by breaking up testing periods into shorter durations over multiple days to ensure all tasks that a participant can complete are tested.

Future work is needed to examine the relationship between skills required for object manipulation and location of damage following the stroke incident to fully translate these findings into clinical practice. Although the present study shows that each of these skills required for object manipulation can be affected in isolation following MCA stroke, to best inform clinical practice, knowing which specific functions are likely preserved in an individual patient will allow for rehabilitation tailored to a patient’s specific pattern of deficits, maximizing reliance on intact systems while minimizing rehabilitative tasks that rely on dysfunctional sensorimotor systems.

## 5. CONCLUSIONS

Taken together, these results demonstrate that each upper-extremity patient shows a unique collection of symptoms, all resulting in deficits in the skillful manipulation of objects. Current bedside diagnostic tests would be well-served by including measurements of each deficit. In particular, the overall effectiveness of rehabilitation interventions could be tied to one or more of these abilities being functional. For example, training that focused on the smooth production and modulation of forces at the fingertips would only have limited success if a patient’s ability to acquire sensorimotor memories is lacking. Ongoing research is examining the functional and anatomical correlates of performance in these tasks while categorizing participants by handedness, lesion location, and lesion lateralization. The information from these future studies may increase our ability to identify each of these deficits, ultimately aiding in future efforts to target rehabilitation regimens to a stroke survivor’s specific pattern of deficits.

## Data Availability

All data produced in the present study are available upon reasonable request to the authors

## Acknowledgements

The authors wish to thank the healthcare professionals at Sanford Health, Avera Health, Rapid City Regional Hospital, and all the participants who volunteered their time to support this research.

## References

Anderson, C., Rajamani, K., Pardo, V., & Adamo, D. E. (2018). Asymmetries in force matching are related to side of stroke in right-handed individuals. Neuroscience Letters, 683, 144–149.

Baugh, L. A., Kao, M., Johansson, R. S., & Flanagan, J. R. (2012). Material Evidence: interaction of well-learned priors and sensorimotor memory when lifting objects. J Neurophysiol, 108, 1262–1269.

Buckingham, G., Cant, J. S., & Goodale, M. A. (2009). Living in a material world: how visual cues to material properties affect the way that we lift objects and perceive their weight. Journal of Neurophysiology, 102, 3111–3118.

Cash, R. F., Isayama, R., Gunraj, C. A., Ni, Z., & Chen, R. (2015). The influence of sensory afferent input on local motor cortical excitatory circuitry in humans. The Journal of physiology, 593, 1667–1684.

Chang, E. C., Flanagan, J. R., & Goodale, M. A. (2008). The intermanual transfer of anticipatory force control in precision grip lifting is not influenced by the perception of weight. Experimental Brain Research, 185, 319–329.

Chouinard, P. A., Leonard, G., & Pauls, T. (2005). Role of the primary motor and dorsal premotor cortices in the anticipation of forces during object lifting. Journal of Neuroscience, 25, 2277–2284.

Cole, K., & Rotella, D. (2002). Old age impairs the use of arbitrary visual cues for predictive control of fingertips during grasp. Experimental Brain Research, 143, 35–41.

Dum, R. P., & Strick, P. L. (1996). Spinal cord terminations of the medial wall motor areas in macaque monkeys. Journal of Neuroscience, 16, 6513–6525.

Edwards, L. L., King, E. M., Buetefisch, C. M., & Borich, M. R. (2019). Putting the “sensory” into sensorimotor control: the role of sensorimotor integration in goal-directed hand movements after stroke. Frontiers in integrative neuroscience, 13, 16.

Eidenmüller, S., Randerath, J., Goldenberg, G., Li, Y., & Hermsdörfer, J. (2014). The impact of unilateral brain damage on anticipatory grip force scaling when lifting everyday objects. Neuropsychologia, 61, 222–234.

Ekstrand, E., Rylander, L., Lexell, J., & Brogårdh, C. (2016). Perceived ability to perform daily hand activities after stroke and associated factors: a cross-sectional study. BMC neurology, 16, 1–9.

Errante, A., Ziccarelli, S., Mingolla, G., & Fogassi, L. (2021). Grasping and Manipulation: Neural Bases and Anatomical Circuitry in Humans. Neuroscience, 458, 203–212.

Flanagan, J. R., & Beltzner, M. A. (2000). Independence of perceptual and sensorimotor predictions in the size-weight illusion. Nat Neurosci, 3, 737–741.

Flanagan, J. R., Bowman, M. C., & Johansson, R. S. (2006). Control strategies in object manipulation tasks. Curr Opin Neurobiol, 16, 650–659.

Flanagan, J. R., King, S., Wolpert, D. M., & Johansson, R. S. (2001). Sensorimotor prediction and memory in object manupulation. Canadian Journal of Experimental Psychology, 55, 87–95.

Folstein, M. F., Folstein, S. E., & McHugh, P. R. (1975). “Mini-mental state”: a practical method for grading the cognitive state of patients for the clinician. Journal of psychiatric research, 12, 189–198.

Gordon, A., Forssberg, H., Johansson, R. S., & Westling, G. (1991). Visual size cues in the programming of manipulative forces during precision grip. Experimental brain research, 83, 477–482.

Gordon, A., Ingvarsson, P. E., & Forssberg, H. (1997). Anticipatory control of manipulative forces in Parkinson’s disease. Experimental neurology, 145, 477–488.

Gordon, A. M., Westling, G., Cole, K. J., & Johansson, R. S. (1993). Memory representations underlying motor commands used during manipulation of common and novel objects. Journal of neurophysiology, 69, 1789–1796.

Grabowski, M., Nordborg, C., & Johansson, B. B. (1991). Sensorimotor performance and rotation correlate to lesion size in right but not left hemisphere brain infarcts in the spontaneously hypertensive rat. Brain Research, 547, 249–257.

Hermsdörfer, J., Hagl, E., Nowak, D. A., & Marquardt, C. (2003). Grip force control during object manipulation in cerebral stroke. Clinical Neurophysiology, 114, 915–929.

Hsu, H.-Y., Ke, C.-W., Kuan, T.-S., Yang, H.-C., Tsai, C.-L., & Kuo, L.-C. (2018). Impacts of sensation, perception, and motor abilities of the ipsilesional upper limb on hand functions in unilateral stroke: quantifications from biomechanical and functional perspectives. PM&R, 10, 146–153.

Jenmalm, P., Schmitz, C., Forssberg, H., & Ehrsson, H. H. (2006). Lighter or heavier than predicted: neural correlates of corrective mechanisms during erroneously programmed lifts. J Neurosci, 26, 9015–9021.

Jo, H. J., Maenza, C., Good, D. C., Huang, X., Park, J., Sainburg, R. L., & Latash, M. L. (2016). Effects of unilateral stroke on multi-finger synergies and their feed-forward adjustments. Neuroscience, 319, 194–205.

Johansson, R. S., & Westling, G. (1988). Coordinated isometric muscle commands adequately and erroneuously programmed for the weight during lifting task with precision grip. Exp Brain Res, 71, 59–71.

Lang, C. E., & Schieber, M. H. (2003). Differential impairment of individuated finger movements in humans after damage to the motor cortex or the corticospinal tract. Journal of Neurophysiology, 90, 1160–1170.

Li, S., Latash, M. L., Yue, G. H., Siemionow, V., & Sahgal, V. (2003). The effects of stroke and age on finger interaction in multi-finger force production tasks. Clinical Neurophysiology, 114, 1646–1655.

Lindberg, P., Ody, C., Feydy, A., & Maier, M. A. (2009). Precision in isometric precision grip force is reduced in middle-aged adults. Exp Brain Res, 193, 213–224.

Lindberg, P. G., Roche, N., Robertson, J., Roby-Brami, A., Bussel, B., & Maier, M. A. (2012). Affected and unaffected quantitative aspects of grip force control in hemiparetic patients after stroke. Brain Research, 1452, 96–107.

Lyle, R. C. (1981). A performance test for assessment of upper limb function in physical rehabilitation treatment and research. Int J Rehabil Res, 4, 483–492.

Mon-Williams, M., & Murray, A. H. (2000). The size of the visual size cue used for programming manipulative forces during precision grip. Exp Brain Res, 135, 405–410.

Naik, S. K., Patten, C., Lodha, N., Coombes, S. A., & Cauraugh, J. H. (2011). Force control deficits in chronic stroke: grip formation and release phases. Experimental Brain Research, 211, 1–15.

Nowak, D. A., Grefkes, C., Dafotakis, M., Küst, J., Karbe, H., & Fink, G. R. (2007). Dexterity is impaired at both hands following unilateral subcortical middle cerebral artery stroke. European Journal of Neuroscience, 25, 3173–3184.

Nowak, D. A., & Hermsdörfer, J. (2003). Selective deficits of grip force control during object manipulation in patients with reduced sensibility of the grasping digits. Neuroscience research, 47, 65–72.

Nowak, D. A., Hermsdorfer, J., & Topka, H. (2003). Deficits of predictive grip force control during object manipulation in acute stroke. J Neurol, 250, 850–860.

Oldfield, R. C. (1971). The assessment and analysis of handedness: the Edinburgh inventory. Neuropsychologia, 9, 97–113.

Quaney, B. M., He, J., Timberlake, G., Dodd, K., & Carr, C. (2010). Visuomotor training improves stroke-related ipsilesional upper extremity impairments. Neurorehabilitation and Neural Repair, 24.

Quaney, B. M., Perera, S., Maketsky, R., Luchies, C. W., & Nudo, R. J. (2005). Impaired grip force modulation in the ipsilesional hand after unilateral middle cerebral artery stroke. Neurorehabilitation and Neural Repair, 19, 338–349.

Quaney, B. M., Rotella, D. L., Peterson, C., & Cole, K. J. (2003). Sensorimotor memory for fingertip forces: evidence for a task-independent motor memory. Journal of Neuroscience, 23, 1981–1986.

Roh, J., Rymer, W. Z., & Beer, R. F. (2015). Evidence for altered upper extremity muscle synergies in chronic stroke survivors with mild and moderate impairment. Frontiers in human neuroscience, 6.

Schaefer, S. Y., Haaland, K. Y., & Sainburg, R. L. (2007). Ipsilesional motor deficits following stroke reflect hemispheric specializations for movement control. Brain, 130, 2146–2158.

Schmitz, C., Jenmalm, P., Ehrsson, H. H., & Forssberg, H. (2005). Brain activity during predictable and unpredictable weight changes when lifting objects. J Neurophysiol, 93, 1498–1509.

Sunderland, A., Bowers, M. P., Sluman, S.-M., Wilcock, D. J., & Ardron, M. E. (1999). Impaired dexterity of the ipsilateral hand after stroke and the relationship to cognitive deficit. Stroke, 30, 949–955.

Velasques, B., Machado, S., Portella, C. E., Silva, J. G., Basile, L. F., Cagy, M., Piedade, R., & Ribeiro, P. (2007). Electrophysiological analysis of a sensorimotor integration task. Neuroscience Letters, 426, 155–159.

Virani, S. S., Alonso, A., Benjamin, E. J., Bittencourt, M. S., Callaway, C. W., Carson, A. P., Chamberlain, A. M., Chang, A. R., Cheng, S., & Delling, F. N. (2020). Heart disease and stroke statistics—2020 update: a report from the American Heart Association. Circulation, 141, e139–e596.

Wolpert, D. M., & Flanagan, J. R. (2001). Motor prediction. Current biology, 11, R729–R732.

Wolpert, D. M., Goodbody, S. J., & Husain, M. (1998). Maintaining internal representations: the role of the human superior parietal lobe. Nature neuroscience, 1, 529–533.

